# Attitudes towards clinical and non-clinical services among individuals who self-harm or attempt suicide: A systematic review

**DOI:** 10.1101/2023.03.15.23287293

**Authors:** Tasnim Uddin, Alexandra Pitman, Gemma Benson, Zeast Kamal, Keith Hawton, Sarah Rowe

## Abstract

The prevalence of self-harm has increased substantially in recent decades. Despite the development of guidelines for better management and prevention of self-harm, service users report that quality of care remains variable. A previous systematic review of research published to June 2006 documented largely negative experiences of clinical services among patients who self-harm. We reviewed research papers published since then until July 2022 to examine contemporary attitudes towards clinical and non-clinical services among individuals who self-harm and their relatives. We identified 29 studies meeting inclusion criteria, all of which were from high- or middle-income countries and were generally of high methodological quality. Our narrative synthesis identified negative attitudes towards clinical management and organisational barriers across services. Generally, more positive attitudes were found towards non-clinical services providing therapeutic contact, such as voluntary sector organisations and social services, than clinical services, such as emergency departments and inpatient units. Views suggested that negative experiences of service provision may perpetuate a cycle of self-harm. Our review suggests that in recent years there has been little improvement in experiences of services for patients who self-harm. These findings should be used to reform clinical guidelines and staff training across clinical services to promote patient-centred and compassionate care and deliver more effective, acceptable and accessible services.

## Introduction

The prevalence of self-harm has increased globally, with evidence of this in countries such as Norway (Tormoen, Myhre, Walby, Groholt, & Rossow, 2020), England (McManus et al., 2019), the United States, China and India (McManus et al., 2019; Muehlenkamp, Claes, Havertape, & Plener, 2012; Tormoen, Myhre, Walby, Groholt, & Rossow, 2020). Psychologically, self-harm is associated with low self-esteem, interpersonal difficulties, and hopelessness (Fox et al., 2015; Hawton, Saunders, & O’Connor, 2012). Physically, self-harm can result in severe scarring, muscle and nerve damage, infection, and premature death (Hawton et al., 2012; Witt et al., 2021b). Self-harm is the strongest predictor of suicide (Carr et al., 2017; Geulayov et al., 2019; Hawton, Zahl, & Weatherall, 2003) with approximately 50% of individuals who die by suicide having previous episodes of self-harm (Fazel & Runeson, 2020; Foster, Gillespie, & McClelland, 1997).

Healthcare services have been criticised over their management of self-harm. Studies demonstrate a high degree of variation in self-harm management across general hospital settings (Arensman et al., 2018; Cooper et al., 2013). For example, the proportion of patient presentations for self-harm receiving psychosocial assessments in emergency departments in England was approximately 58% although it ranged by hospital from 28% to 91%, (Cooper et al., 2015) despite this being recommended practice for self-harm presentations (NICE, 2022). There is also evidence to support the effectiveness of interventions in preventing repeat self-harm or suicide following a first episode (Witt et al., 2021a, 2021b). Rates of readmission to psychiatric inpatient care for self-harm are highest in the following year, with one third of these occurring in the first month after discharge (Gunnell et al., 2008). Despite this, national guidelines for the short-term management of self-harm have been found to be implemented by healthcare professionals in less than half of the encounters they have with patients (Leather et al., 2020). Together, this evidence highlights a need for standardised and improved care.

Eliciting patients’ attitudes towards services providing interventions for self-harm are essential as they identify barriers to service delivery and influence treatment engagement (Ribeiro Coimbra & Noakes, 2022). The ‘*Interpersonal cycle of reinforcement of* self *injury’* (Rayner, Allen, & Johnson, 2005) posits that patients’ experiences of stigmatising attitudes from staff and negative therapeutic relationships can feed into negative cognitions about themselves, which can lead to treatment disengagement. Understanding patients’ experiences of services therefore enables identification of key areas of improvement to enhance treatment adherence and improve outcomes (N. Kapur et al., 2013; Rayner et al., 2005; Ribeiro Coimbra & Noakes, 2022).

A systematic review of patients’ attitudes towards clinical services following self-harm published in 2009 identified predominantly negative perceptions, including poor communication between patients and staff, limited staff knowledge of self-harm and poor therapeutic relationships (Taylor, Hawton, Fortune, & Kapur, 2009). Many patients suggested a need for improvements in psychosocial assessment, referral pathways and access to after-care. As that review was completed over a decade ago and focussed only on clinical services, an update of the literature is needed to reflect contemporary practice, widening the scope to the full range of services currently available to people who self-harm. The present systematic review aimed to examine attitudes of patients and their families towards clinical and non-clinical self-harm services from research published since the final search date of the previous review (July 2006). We also aimed to compare patients’ experiences of clinical and non-clinical services, defining clinical services as those provided by public or private healthcare providers (primarily consisting of clinicians), and non-clinical services as charitable and voluntary sector organisations, social services, and faith-based organisations.

## Method

Our review followed the Preferred Reporting Items for Systematic Reviews and Meta-analyses (PRISMA) guidelines (Moher, Liberati, Tetzlaff, Altman, & Group, 2009). We pre-registered the review protocol with PROSPERO (CRD42021264789).

### Search Strategy

As our review represented an update of a previous systematic review (Taylor et al., 2009), we replicated their methodology but expanded our search terms to include clinical and non-clinical services, and updated terminology (supplementary materials: S1).

We searched seven electronic databases (EMBASE, MEDLINE, PsycINFO, Global Health, AMED, HMIC and CINAHL). We also searched Google Scholar and OpenGrey for grey literature. Papers were limited to those in English language and published from July 2006 as the previous review included papers up until June 2006 (Taylor et al., 2009). The initial search was conducted in July 2021 and the final search was conducted on 1^st^ July 2022. The reference lists of included studies were hand-searched to identify further eligible studies.

### Inclusion and exclusion criteria

We included published and unpublished primary research studies capturing the experiences or attitudes towards services of people who self-harm. Eligible studies were those that included participants with at least one episode of self-harm, irrespective of suicidal intent. Studies were excluded if participants experienced attempts of assisted suicide, euthanasia attempts or experienced harm without explicit intent (e.g., accidental overdose). We also included studies capturing the attitudes of carers and relatives of individuals who self-harmed. Studies were included if participants received any medical or psychosocial intervention for their self-harm episode from clinical services (primary or secondary healthcare) or non-clinical services (services outside of healthcare settings including but not limited to social, voluntary sector or faith-based services). In order to maximise the evidence, qualitative, quantitative and mixed methods studies were included, as was the case in the previous review (Taylor et al., 2009). Secondary analyses of data and systematic reviews were excluded.

### Study Selection

Search results were exported into Covidence (Covidence systematic review software, 2021) and de-duplicated. All titles and abstracts were first screened by one reviewer (TU). Full text articles of eligible studies were then screened by a second independent reviewer (ZK or GB) using the predetermined inclusion and exclusion criteria. Any disagreements were resolved through discussions with a third reviewer (SR).

### Data extraction

A data extraction table was used to extract information on authors, publication year, country of origin, sample size, sample characteristics (i.e., demographic information), type of self-harm behaviours, type of services and interventions, methodology, measures of attitudes and relevant quantitative and/or qualitative findings. All data were extracted by one reviewer (TU) and independently verified by a second reviewer (ZK or GB).

### Quality assessment

The quality of included papers was assessed using the Mixed Methods Appraisal Tool (MMAT) (Hong et al., 2018) by one reviewer (TU) and was independently verified by a second reviewer (ZK or GB). The MMAT has previously been validated for use in systematic reviews and was selected as it is designed to appraise a variety of study designs (Hong et al., 2018). Calculating an overall quality score is discouraged when using the MMAT, therefore, we reported scores for each criterion. There were high levels of agreement between the reviewers, with only one paper requiring discussion. All papers were given equal value in terms of contributing to the findings.

### Data synthesis

We summarised quantitative and qualitative findings using a narrative synthesis approach as we anticipated a wide variety of study designs, sample populations and measures and therefore substantial heterogeneity of findings. We used validated guidelines for narrative syntheses from the Economic and Social Research Council framework to follow established practice (Popay, 2006).

One researcher (TU) first grouped studies by methodology, setting and population, tabulating key findings relevant to attitudes towards services using these categories. Team discussions were used to agree these categories. Findings were then compared across studies to categorise similarities and differences in attitudes by setting and population, and to identify meaningful higher-level constructs (Popay, 2006). The final constructs were synthesised following critical discussion with the wider team until complete agreement on structure and content was reached.

Finally, we sought the perspective of an individual with lived experience of accessing self-harm services to help us interpret findings.

## Results

### Study selection

The initial search identified 9,443 studies, which was reduced to 6,028 studies following deduplication (Figure 1). Full text screening was completed on 142 papers with 26 studies were deemed eligible and included in the review. Three further studies were identified from hand-searching the reference list of these included articles. A total of 29 studies were included.

**Figure 1.**
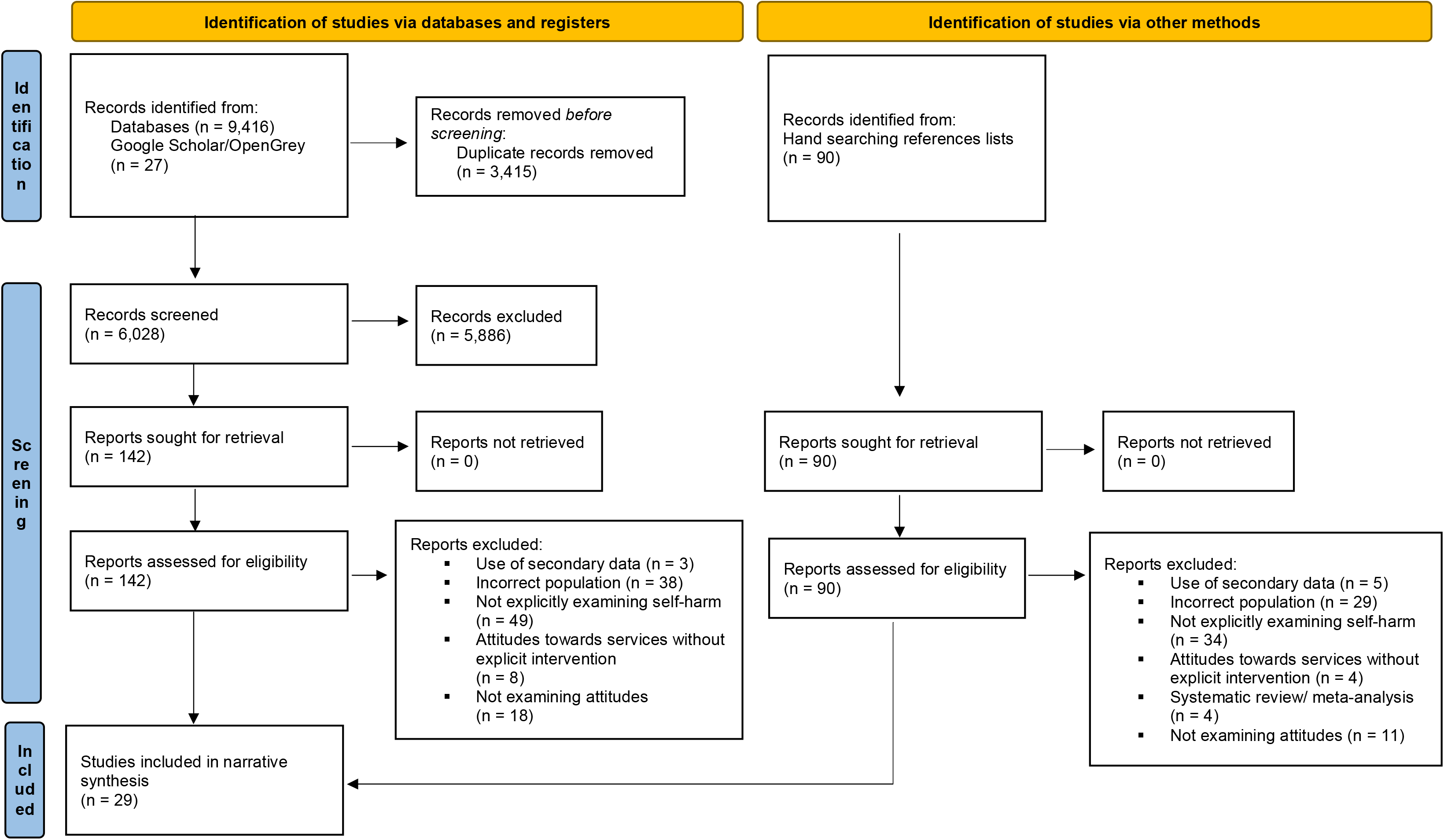
PRISMA flowchart describing the study selection process.

### Study characteristics

Study characteristics of the included studies are summarised in Table 1. Studies were published between 2007 to 2022, all in high- and middle-income countries: these included 11 studies from the UK, four from Sweden, two from Canada, two from China, two from Norway, two from the USA, one from Australia, one from Belgium, one from Finland, one from Ireland, one from Portugal, and one from South Africa. The gender profiles of participants were reported in 25 studies. While one study included only female participants (Lindkvist et al., 2021) and one included only male participants (Hassett & Isbister, 2017), all other studies included a mix of female and male participants, with four studies including transgender, non-binary or other genders. Only three studies reported on participants’ ethnicity, all of which included exclusively or majority White participants.

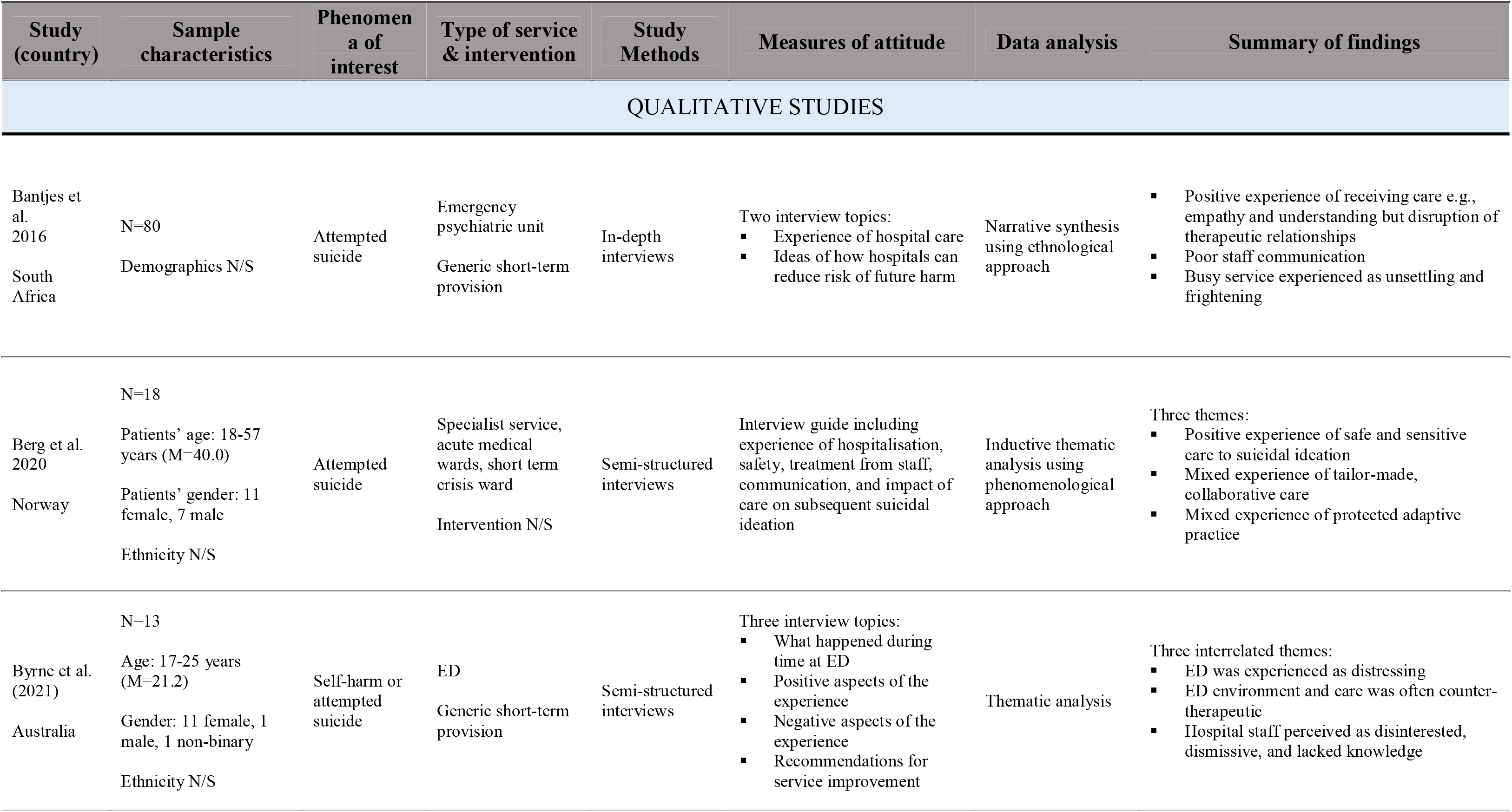
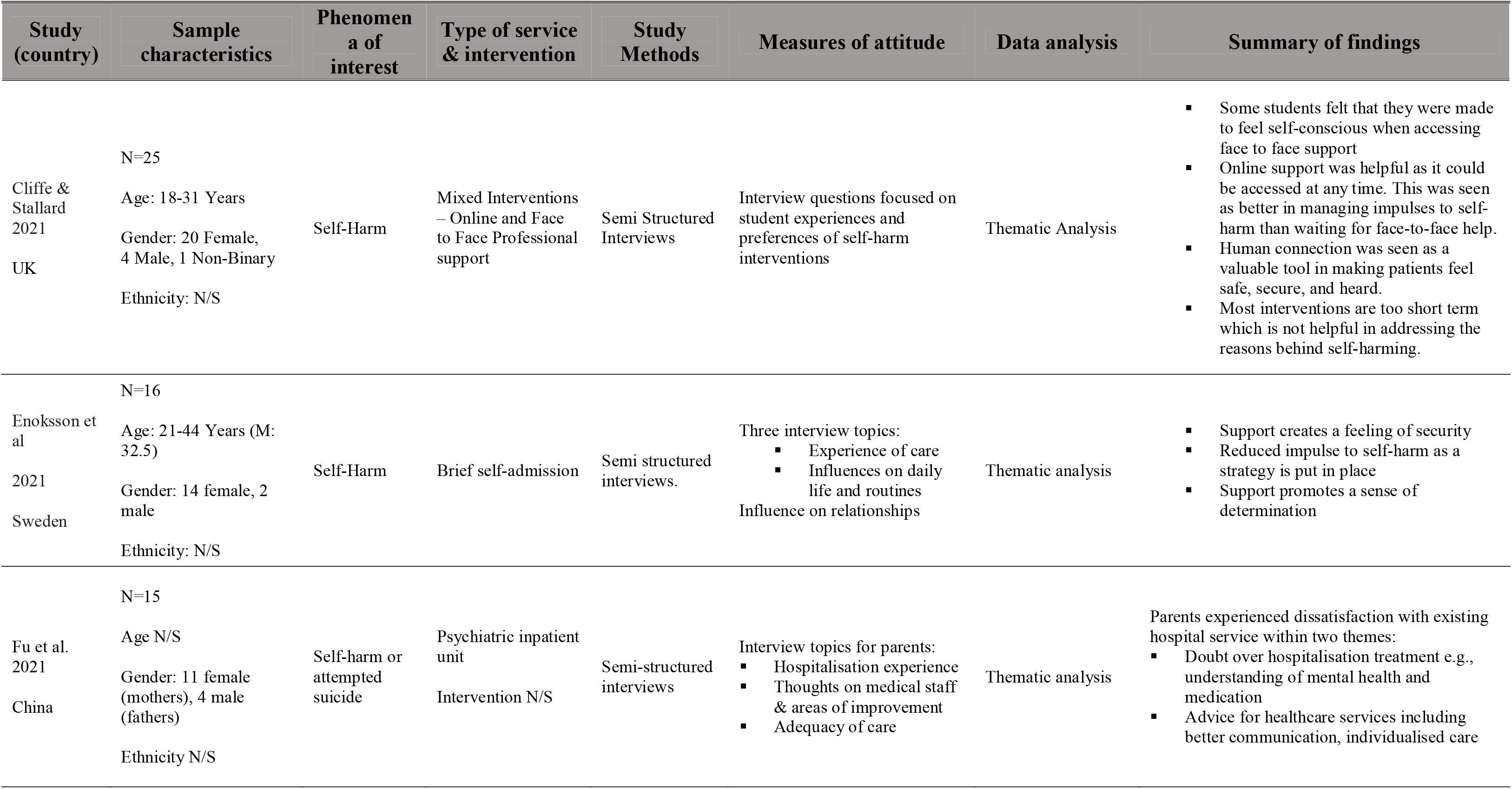
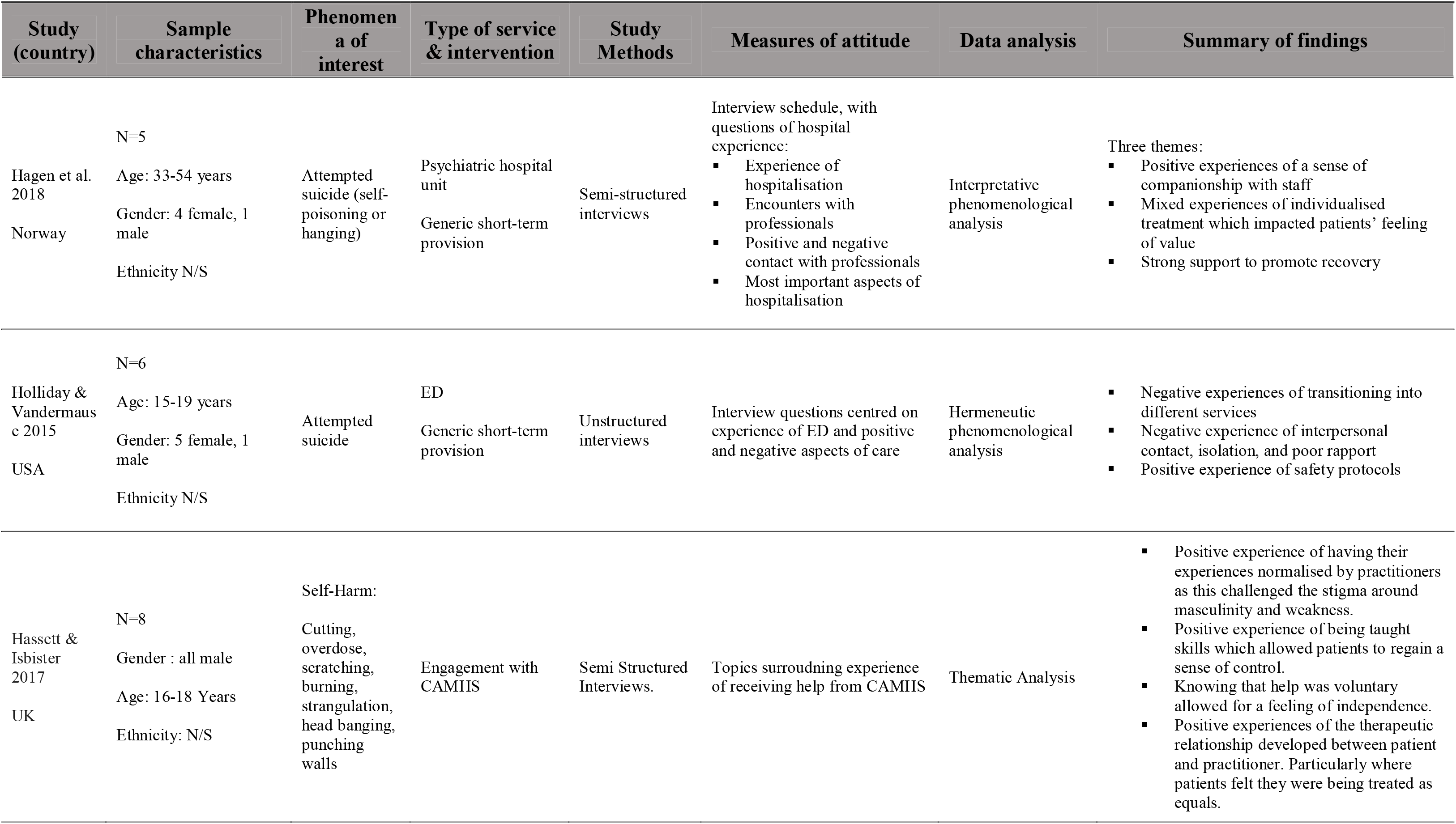
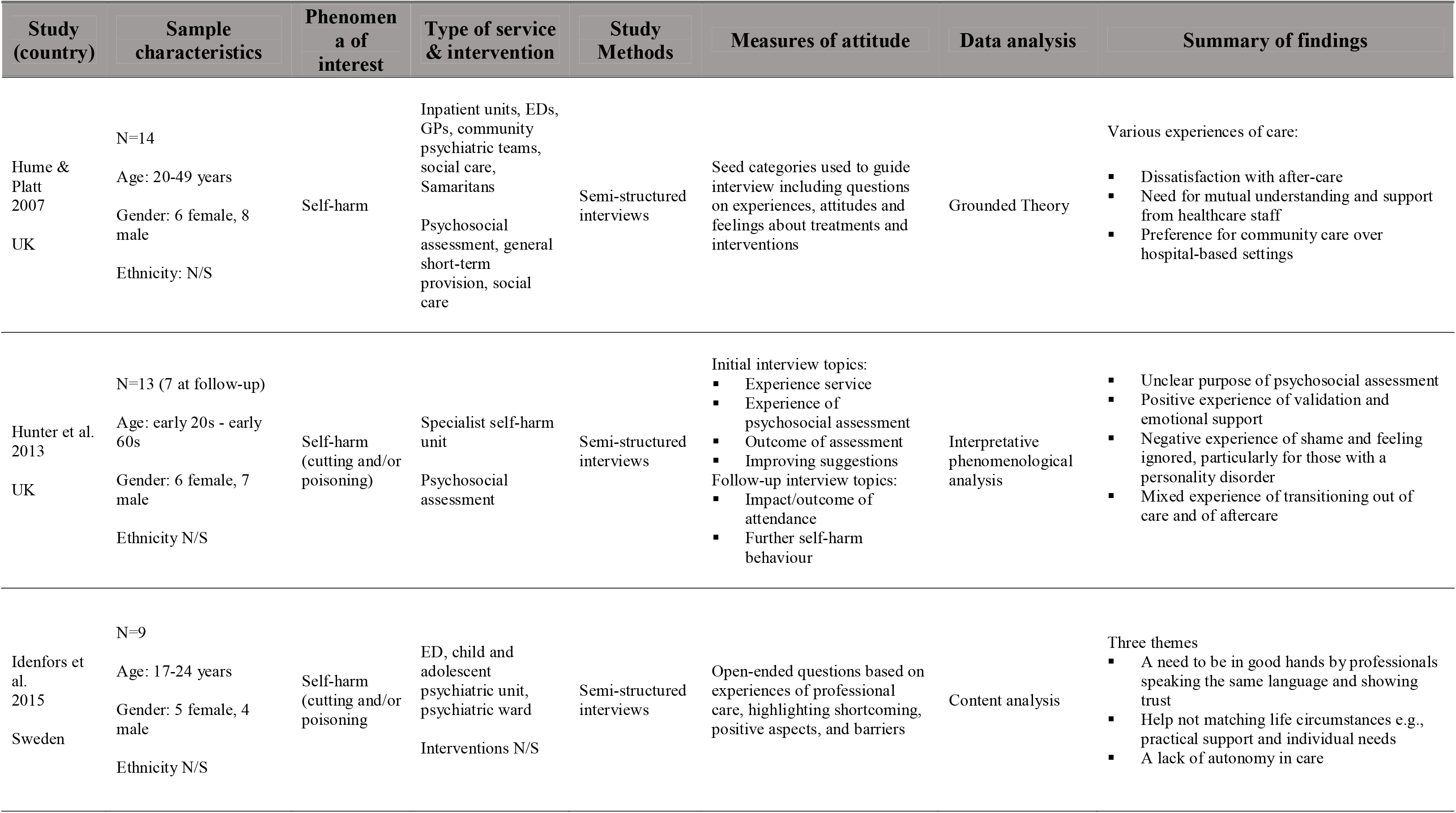
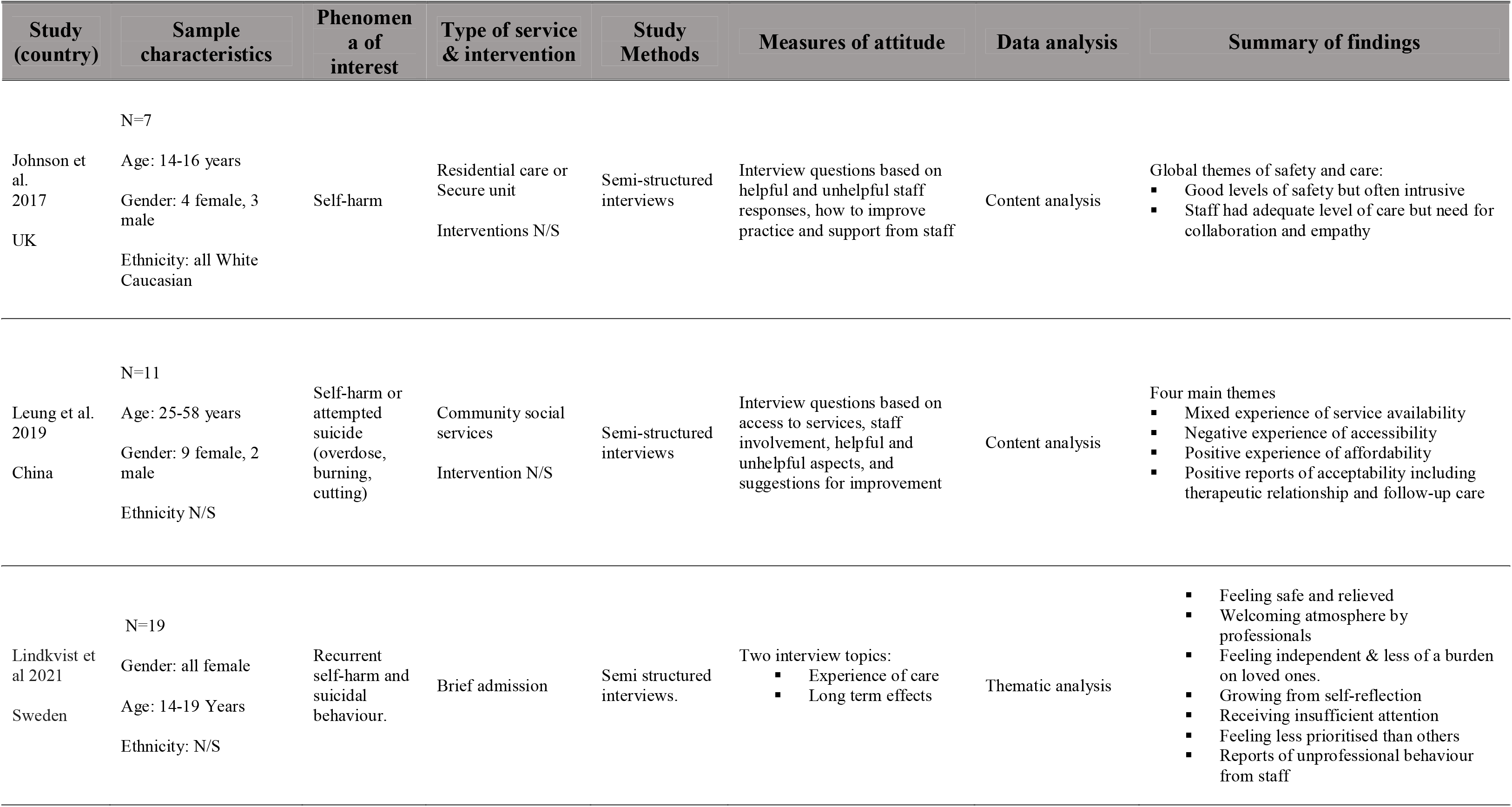
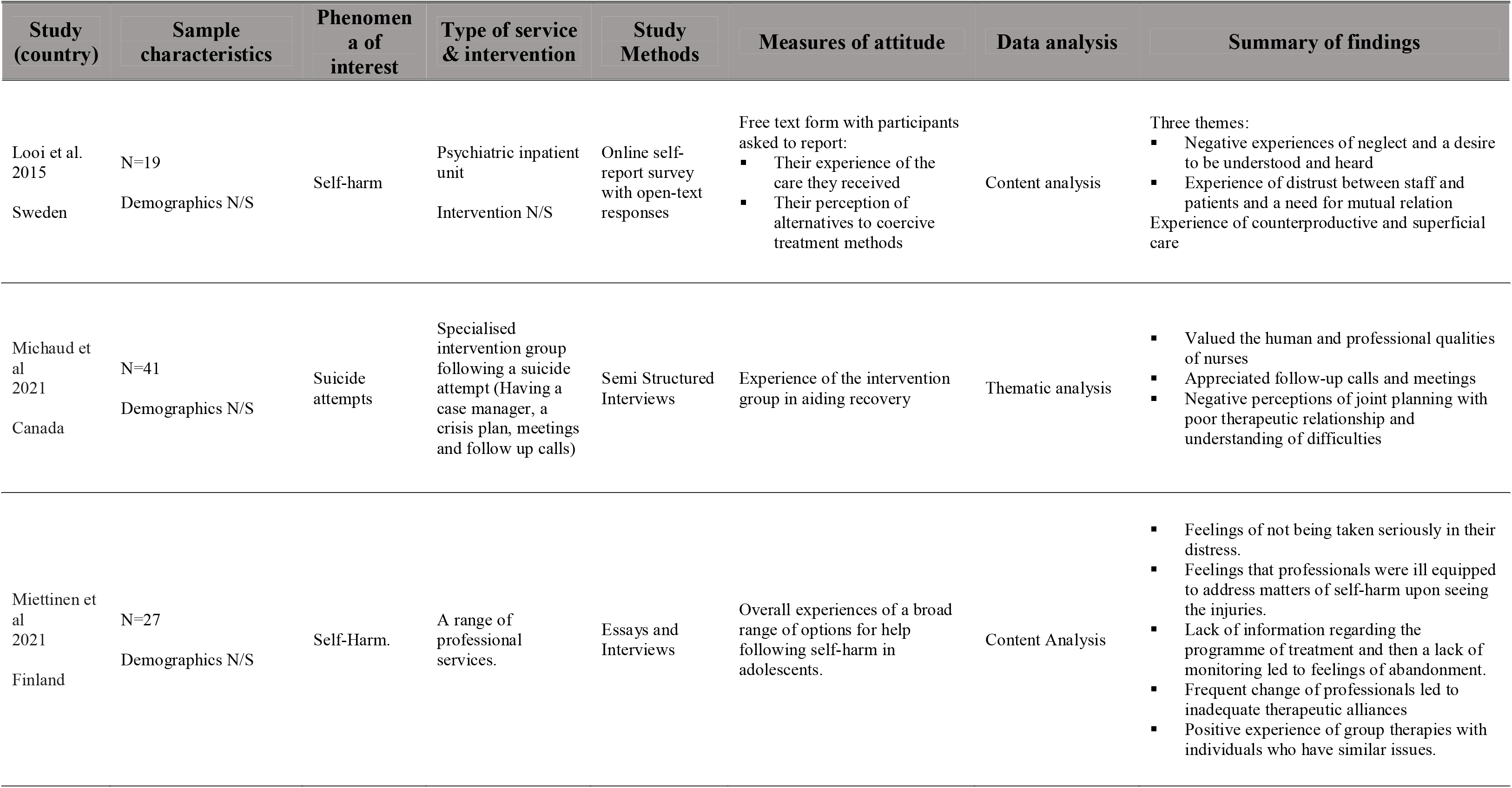
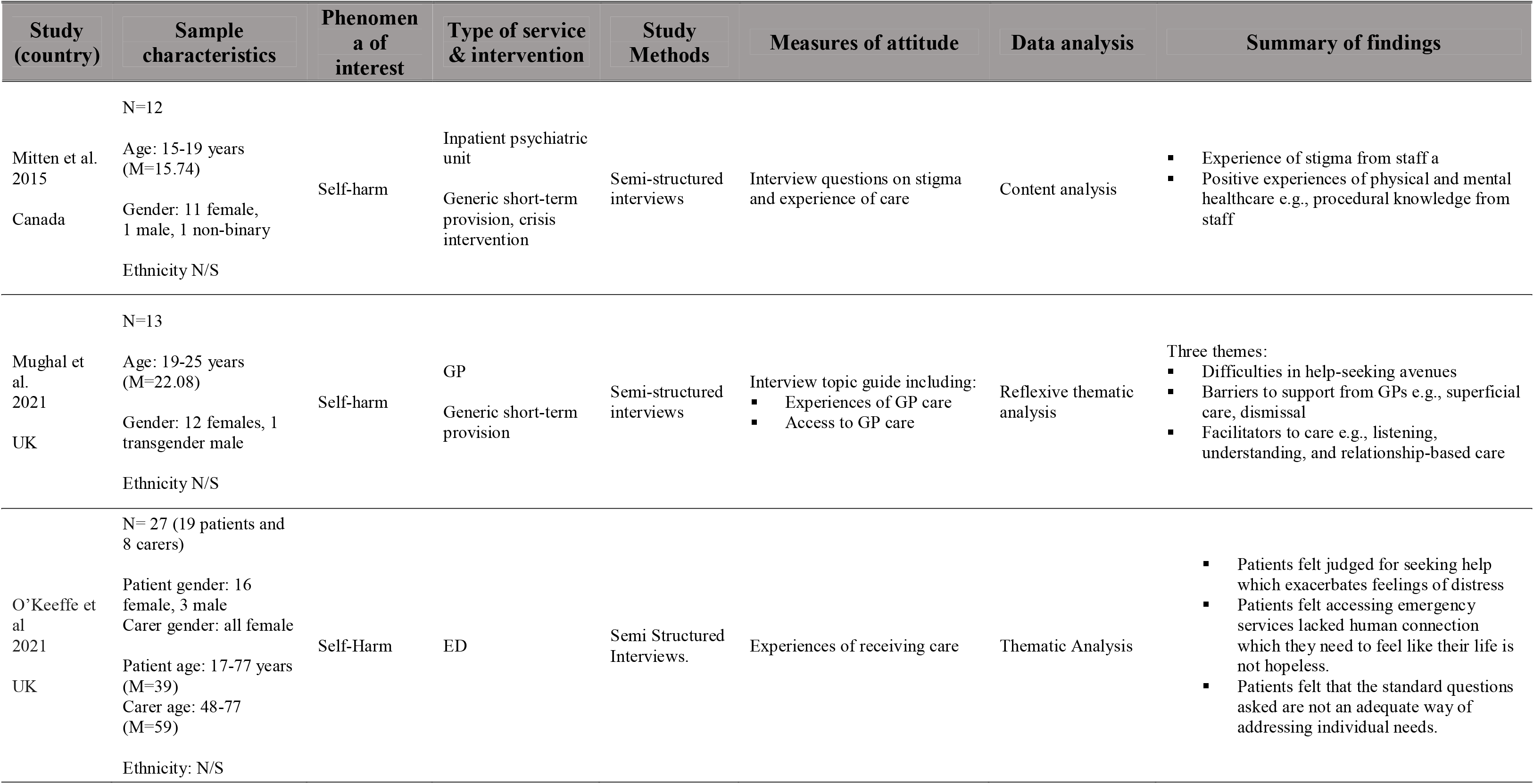
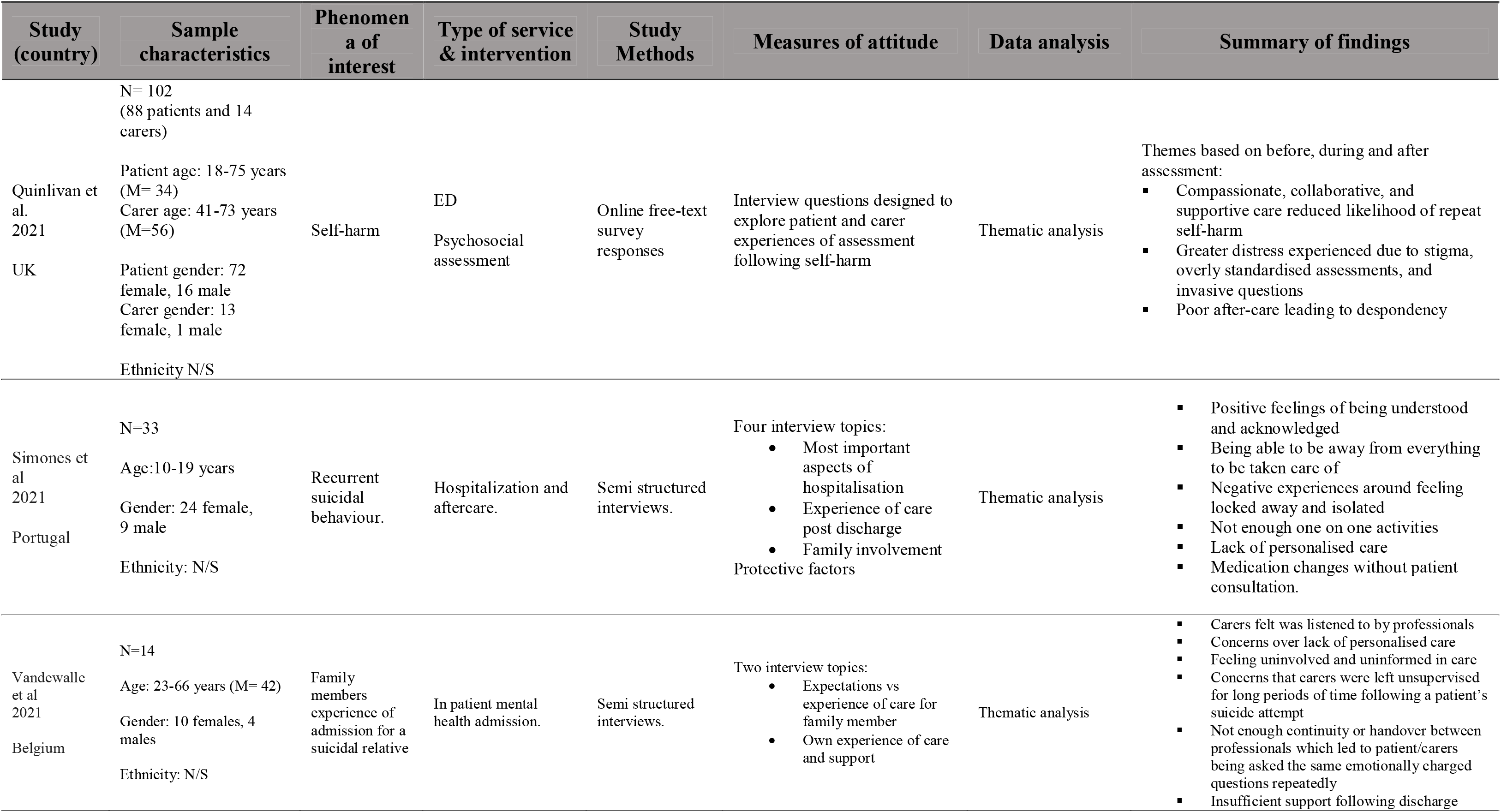
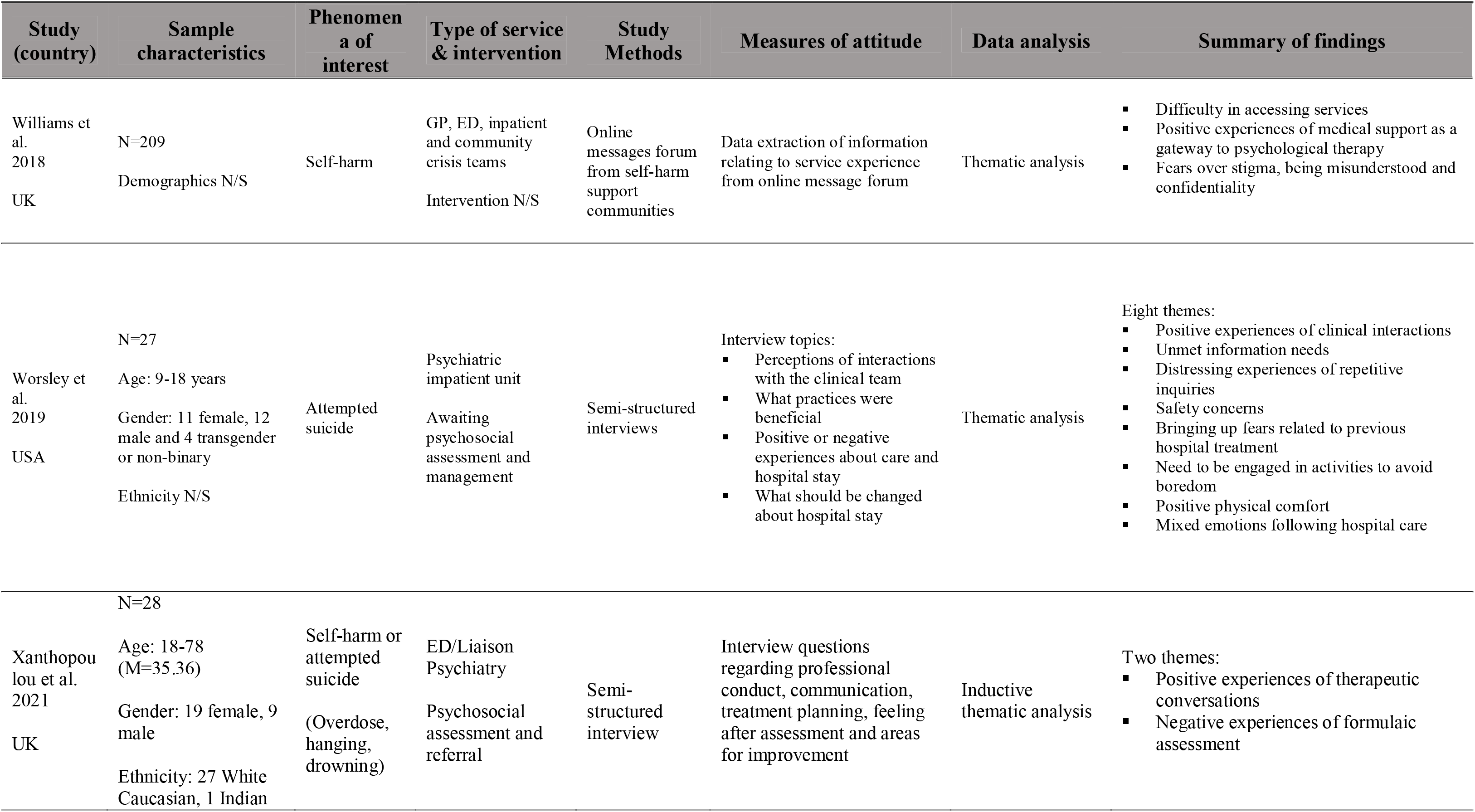
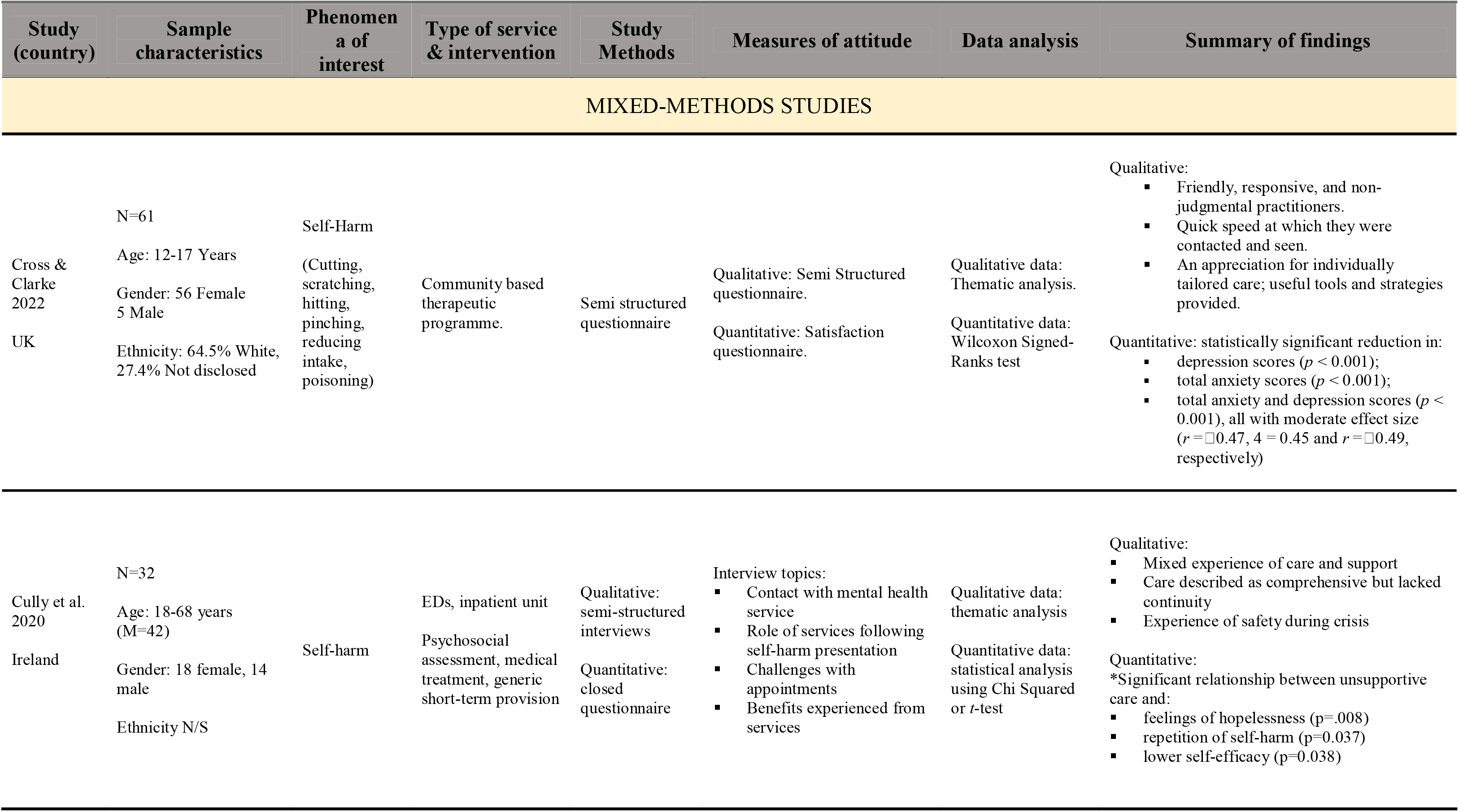
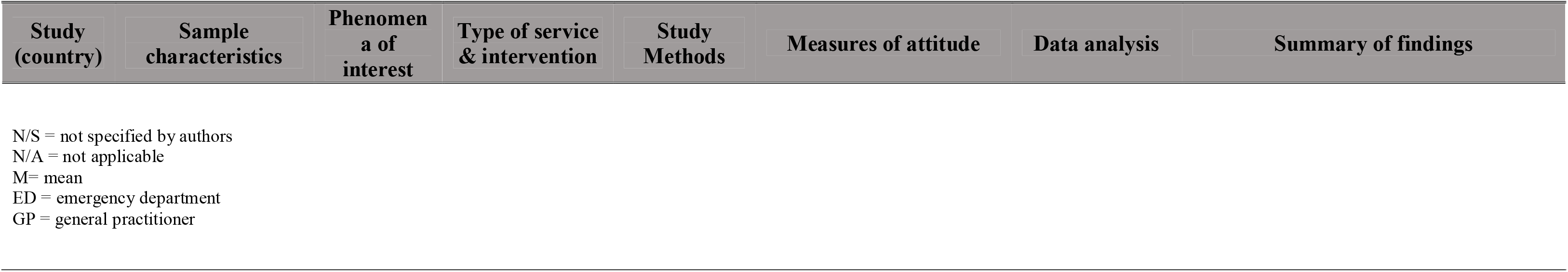

The studies examined attitudes of patients/carers following a patient’s presentation for self-harm (n=16), attempted suicide (n=8) or a mixture of self-harm and attempted suicide (n=5). Studies examined patients’ attitudes solely (n=24), relatives’ attitudes solely (n=2) or both patients’ and relatives’ attitudes (n=3). Studies exclusively examined one type of service (n=18) or a combination of services (n=11). The clinical services included in studies were psychiatric/inpatient units (n=12), emergency departments (EDs; n=10), primary care (n=4), secure units (n=1), crisis wards/brief admission units (n=3), community-based psychiatric teams (n=3), community-based crisis care (n=2), specialist psychiatric wards (n=1), acute medical wards (n=1) and Child and Adolescent Mental Health Services (n=1). The non-clinical services included in studies were voluntary sector community-based programmes (n=1), social services (n=2) or a voluntary sector helpline (Samaritans; n=1). Based on these categories we made a team decision to group findings by clinical versus non-clinical services.

### Quality assessment

Quality assessment ratings for the studies are presented in Table 2a and 2b. We judged 25 of the 27 qualitative studies to be of high methodological quality. Both the mixed-methods studies were assessed to be of moderate risk of bias.

**Table 2a.**
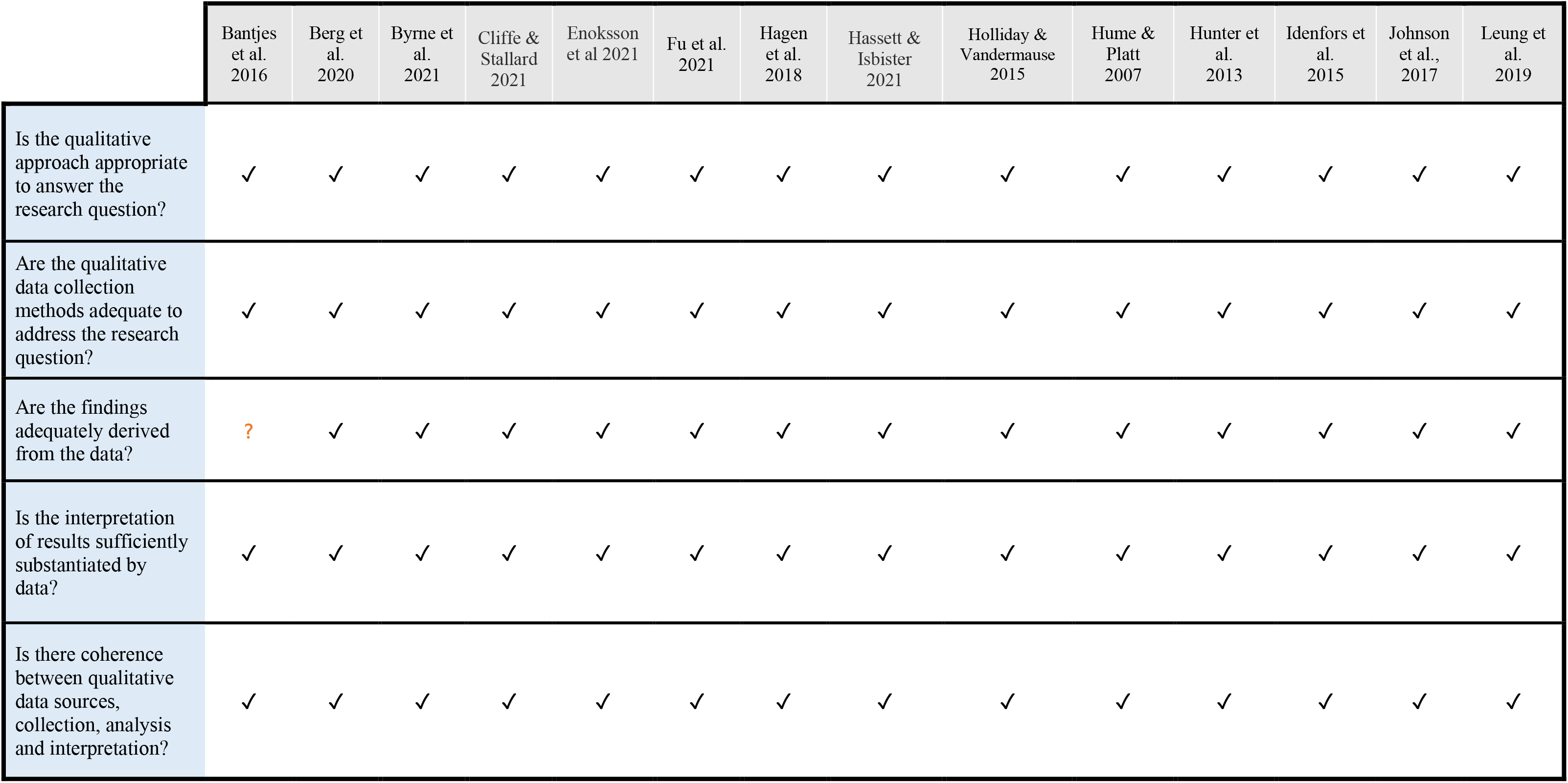
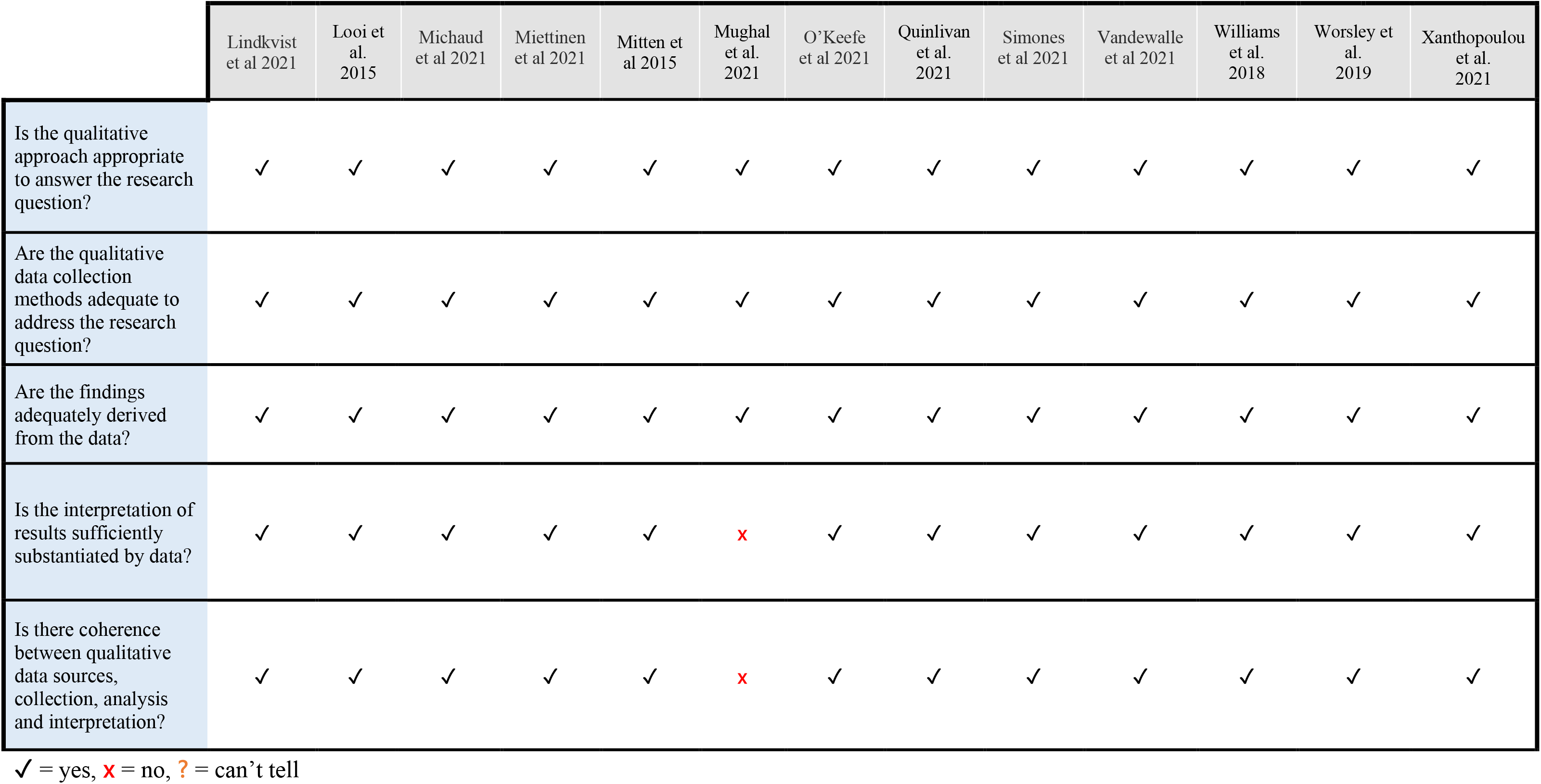
Quality assessment ratings for qualitative studies using the MMAT.

**Table 2b.**
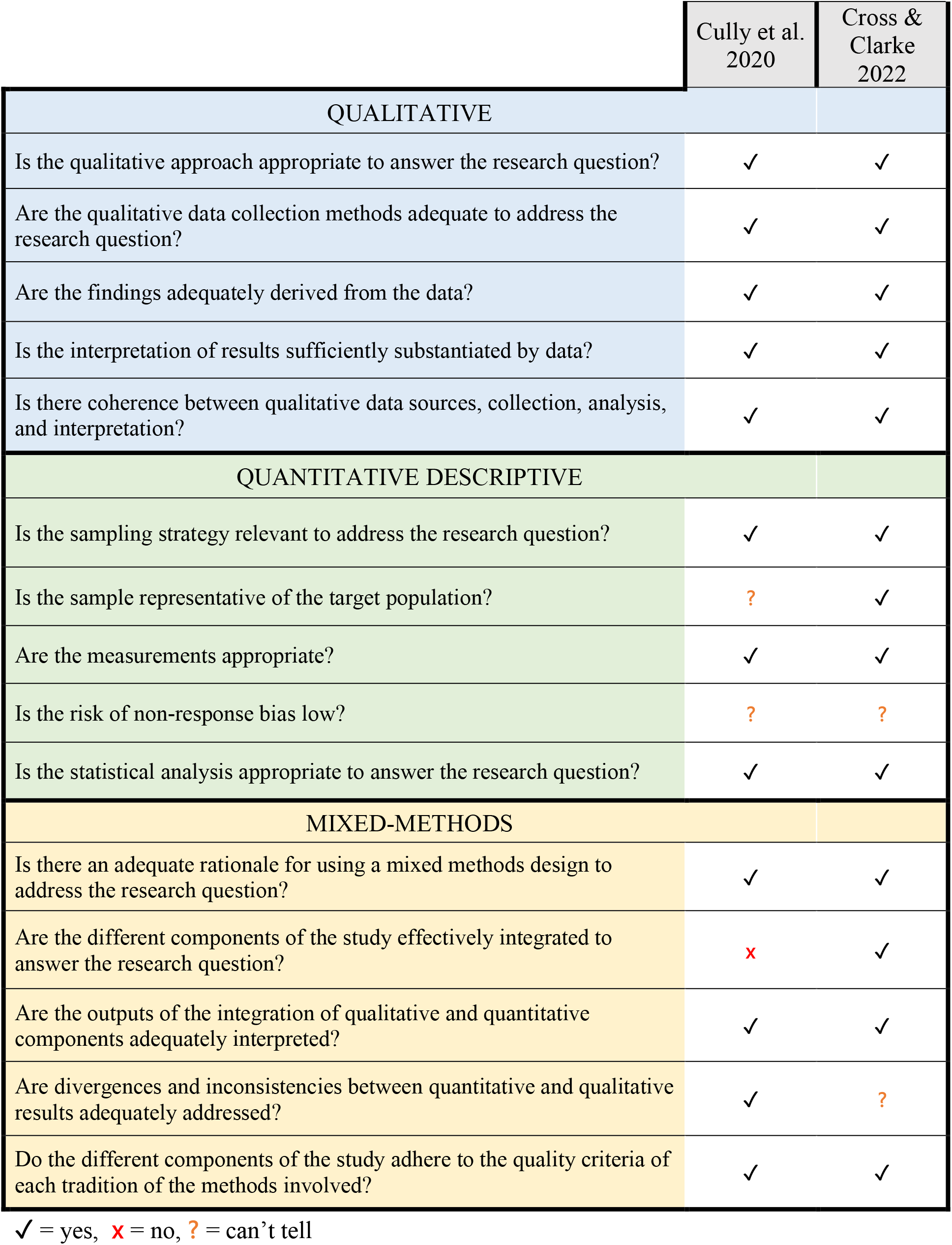
Quality assessment ratings for mixed-methods study using the MMAT.

### Attitudes towards services from individuals who self-harm and their relatives

Our narrative synthesis of studies resulted in the development of four overarching constructs: *staff attitudes, therapeutic contact, clinical management, and organisational barriers*.

#### Staff attitudes

##### Professional stigma

The stigmatising attitudes of professionals were reported in nine studies that examined clinical services. Across EDs and inpatient units, patients experienced negative judgements, service gate-keeping or belittling comments regarding their injuries (Mitten, Preyde, Lewis, Vanderkooy, & Heintzman, 2016; Quinlivan et al., 2021; Williams, Nielsen, & Coulson, 2020). Five studies reported a perception that professional stigma acted as a barrier to disclosure, with shame and fear impairing disclosure within psychosocial assessments and when help-seeking (Byrne et al., 2021; Hunter, Chantler, Kapur, & Cooper, 2013; Mitten et al., 2016; O’Keeffe, Suzuki, Ryan, Hunter, & McCabe, 2021; Xanthopoulou, Ryan, Lomas, & McCabe, 2022). Patients reported how their own low self-esteem and self-blame were reinforced by professionals’ stigmatising attitudes (Byrne et al., 2021; Quinlivan et al., 2021; Vandewalle et al., 2019).

Experiences of professionals’ stigmatising attitudes varied between clinical and non-clinical services, with the latter preferred for being more accepting. In one study, patients showed preferences for social services and voluntary sector organisations over hospital services, with the former described as more supportive and having the potential to build long-term relationships with patients (Hume & Platt, 2007). In one community-based programme, staff (voluntary sector youth workers) were described as non-judgemental and friendly, reducing any shame felt by patients (Cross & Clarke, 2022).

Two studies set in clinical services described perceptions of stigma surrounding mental health diagnoses. Patients highlighted how professionals’ interest and compassion diminished after disclosure of a diagnosis of a ‘personality disorder’, with labels of “time-waster” and “attention-seeker” applied (Quinlivan et al., 2021). Whilst one UK-based qualitative study reported experiences of staff withdrawal and rushed assessments’(Hunter et al., 2013), another UK-based qualitative study reported perceptions of psychiatric diagnoses being wrongfully used by professionals to minimise the severity of a patient’s self-harm on the basis it was expected or normalised (Quinlivan et al., 2021).

##### Minimisation of distress

A tendency to minimise patients’ distress was reported in eight studies. Across EDs, GPs and inpatient units, staff were described as uninterested and dismissive of physical and psychosocial distress (Ejneborn Looi, Engström, & Sävenstedt, 2015; Hagen, Knizek, & Hjelmeland, 2018; Lindkvist et al., 2021; Mughal, Dikomitis, Babatunde, & Chew-Graham, 2021; Xanthopoulou et al., 2022). Three studies set in clinical services reported experiences of staff prioritising cases that they perceived as more ‘serious’ and patients whose injuries were not self-inflicted, further demonstrating professional discrimination (Ejneborn Looi et al., 2015; Fu et al., 2021; Hagen et al., 2018). Minimisation also resulted in care being withheld; patients were told that pain medication and medical treatments were unnecessary, with staff making comments about a ‘waste’ of beds and resources (Byrne et al., 2021; Hagen et al., 2018; Quinlivan et al., 2021). Minimisation led to patients viewing services as “cold” and “robotic”, only responding if a ‘threshold’ of seriousness was met (Byrne et al., 2021).

#### Therapeutic contact

##### Staff-patient relationship

Twenty-one studies presented data describing relationships with staff. Within non-clinical services (social services and voluntary sector services), patients generally described a strong rapport between themselves and staff, based on mutual understanding, non-judgemental care, and trust (Cross & Clarke, 2022; Hume & Platt, 2007; Leung, Chow, Ip, & Yip, 2019). However, experiences within clinical services were variable. Studies reporting positive experiences highlighted genuine and sensitive contact as well as mutual understanding to empower patients and encourage them to collaboratively explore their distress (Cliffe & Stallard, 2022; Enoksson, Hultsjo, Wardig, & Stromberg, 2022; Hagen et al., 2018; Hassett & Isbister, 2017; Lindkvist et al., 2021; Michaud, Dorogi, Gilbert, & Bourquin, 2021; Xanthopoulou et al., 2022). This rapport allowed staff to respond to patients’ individual needs for more effective care, such as reacting to fluctuations in suicidality, distress, and instability (Berg, Rortveit, Walby, & Aase, 2020; Quinlivan et al., 2021; Worsley, Barrios, Shuter, Pettit, & Doupnik, 2019). Positive rapport allowed patients to feel acknowledged as human beings, which instilled hope for recovery (Hagen et al., 2018; Worsley et al., 2019).

However, these findings contrasted with reports of superficial contact within clinical services (EDs, inpatient and psychiatric units), whereby patients perceived staff as being disconnected and making little effort to engage with their individual experiences (Bantjes et al., 2017; Cully, Leahy, Shiely, & Arensman, 2022; Idenfors, Kullgren, & Salander Renberg, 2015; Miettinen, Kaunonen, Kylma, Rissanen, & Aho, 2021; O’Keeffe et al., 2021; Quinlivan et al., 2021; Simoes, Dos Santos, & Martinho, 2021; Worsley et al., 2019). Three studies highlighted how perceived mistrust from clinical staff impaired patients’ feeling of safety and willingness to engage (Ejneborn Looi et al., 2015; Holliday & Vandermause, 2015; Hume & Platt, 2007). In one quantitative study, perceptions of unsupportive care were significantly associated with repeat self-harm (Cully et al., 2022).

##### Relationships with relatives

Relatives of patients also reported negative experiences within EDs and inpatient units, with four studies highlighting their observations of poor communication from staff. Relatives were often excluded from discussions about patients’ care, felt inadequately informed about prognosis and had their concerns dismissed (Fu et al., 2021; Quinlivan et al., 2021; Vandewalle et al., 2019). Relatives experienced superficial and judgemental staff contact, particularly during sensitive discussions about the patients’ care and self-harm. This led to a lack of confidence in staff and doubts over the quality of care (O’Keeffe et al., 2021; Quinlivan et al., 2021).

#### Clinical management

##### Psychosocial assessment

Attitudes towards psychosocial assessments within clinical settings were reported in eleven studies. Assessments were described as superficial, rushed, and formulaic, where generic tick-box questions denied opportunities to explore individual experiences and psychosocial difficulties (Berg et al., 2020; Byrne et al., 2021; Quinlivan et al., 2021; Simoes et al., 2021). While good staff knowledge of psychosocial assessment protocols was reported in EDs and psychiatric wards, knowledge about mental health in those settings was seen as insufficient, with patients recommending staff training to help them better assess the context for and severity of a patient’s suicidality (Hagen et al., 2018; Holliday & Vandermause, 2015).

Across clinical services, patients and relatives reported a lack of involvement in treatment planning, with unnecessary repetition of questions leading them to believe that staff did not listen or understand their individual experiences (Fu et al., 2021; Quinlivan et al., 2021). However, care was positively experienced when staff were sensitive to patients’ emotional distress when completing an assessment, collaboratively explored the factors leading to self-harm and involved patients in treatment decisions (Johnson, Ferguson, & Copley, 2017; Michaud et al., 2021; Worsley et al., 2019; Xanthopoulou et al., 2022).

##### Use of restrictions and coercive care

Eleven studies reported variable attitudes towards coercive care in clinical services. In five studies, patients and relatives described the benefits of restrictions and removal of potentially lethal objects to protect against further self-harm (Berg et al., 2020; Cully et al., 2022; Hassett & Isbister, 2017; Idenfors et al., 2015; Vandewalle et al., 2019). Many patients experienced EDs and inpatient wards as ‘safe havens’ that removed them from distressing environments (e.g., difficult home dynamics) meaning patients could effectively shift focus towards recovery (Cully et al., 2022; Worsley et al., 2019). Brief admissions empowered some patients as they felt they were given more control over care through joint decision making (Enoksson et al., 2022; Lindkvist et al., 2021). However, other clinical services were experienced more negatively as patients reported feeling disempowered by restrictions (Quinlivan et al., 2021; Simoes et al., 2021). In light of this, patients and relatives expressed the importance of communicative practice when imposing restrictions: where staff in EDs explained the rationale behind restrictions and used collaborative assessments, these mitigated feelings of anxiety and disempowerment (Quinlivan et al., 2021).

##### Discharge and aftercare

Negative experiences of discharge following an assessment for self-harm in clinical services were reported across 12 studies. Studies reported how patients felt ill-prepared and unsafe at discharge where feelings of abandonment diminished their trust in clinical services and triggered repeat self-harm (Berg et al., 2020; Byrne et al., 2021; Hume & Platt, 2007; Idenfors et al., 2015; Xanthopoulou et al., 2022).

Regarding aftercare, some patients were not contacted by services at all, whilst other patients faced long waiting times (Hunter et al., 2013; Quinlivan et al., 2021). Those who did receive follow-up care were often disappointed due to its brief length, low number of appointments given, and prioritisation of discussions about medication over psychology (Cully et al., 2022; Holliday & Vandermause, 2015; Miettinen et al., 2021; Quinlivan et al., 2021). However, two studies of clinical services investigating experiences of patients on brief admission units described positive accounts of detailed discharge plans and safety planning which provided patients with a sense of security (Enoksson et al., 2022; Lindkvist et al., 2021). Greater control over their care meant patients could readjust back into society comfortably (Enoksson et al., 2022; Lindkvist et al., 2021).

##### Psychotropic medication

Seven studies reported on attitudes towards medication administration after self-harm, all of which were within clinical services: EDs, inpatient units and community-based psychiatric care. While medication was seen as helpful, staff were perceived to focus more often on describing benefits whilst tending to minimise information on side-effects and risks (Ejneborn Looi et al., 2015; Idenfors et al., 2015). Changes in medication without follow-up consultations from staff led patients to view services as negligent (Hagen et al., 2018; Simoes et al., 2021). Patients and relatives reported that medication was often administered without adjunctive psychological interventions, which they experienced as avoiding problems rather than an effective resolution (Fu et al., 2021; Hunter et al., 2013; Vandewalle et al., 2019).

#### Organisational barriers

##### Waiting times

Nine studies described negative experiences in clinical services of long waiting times across services. For EDs, inpatient and crisis management teams, lengthy waiting times for a psychosocial assessment led to feelings of anxiety, particularly when in busy and loud environments (Bantjes et al., 2017; Byrne et al., 2021; Miettinen et al., 2021; Quinlivan et al., 2021; Williams et al., 2020). Patients and relatives also received little communication regarding the purpose of the wait, reasons for delays and progress (Cully et al., 2022; Vandewalle et al., 2019). Beyond the ED, there were also experiences of long waiting times for aftercare following an initial assessment (Byrne et al., 2021; Miettinen et al., 2021).

In non-clinical settings, experiences were variable. One community-based programme had an average waiting time of 1.7 days between assessment and referral contact, which patients cited as a key reason for high satisfaction (Cross & Clarke, 2022). However, long waiting times within social services were found to heighten patient anxiety (Leung et al., 2019).

##### Access to care

Nine studies reported variable access to care across clinical services. EDs, inpatient units and brief admission units were reported as having a lack of beds and staff, which patients felt contributed to excessive waiting times, inappropriate transfers, and premature discharges (Byrne et al., 2021; Enoksson et al., 2022; Johnson et al., 2017; Miettinen et al., 2021). For brief admission, some patients felt the care was less specialised compared to what they would receive in EDs and wanted more options for psychological support (Lindkvist et al., 2021). However, others felt that they could call on staff freely within brief admission wards and also a sense of predictability and safety, unlike in busy and intense EDs (Lindkvist et al., 2021).

Many patients were unaware of what non-clinical services were available to them and felt that they should be better integrated with clinical services for more accessible care following discharge (Cross & Clarke, 2022; Leung et al., 2019). For social and voluntary services, they suggested extended services hours, telephone/digital appointments, and better staffing to improve accessibility (Idenfors et al., 2015; Leung et al., 2019; Williams et al., 2020).

## Discussion

### Main findings

This systematic review of 29 studies examined attitudes towards clinical and non-clinical services of individuals who self-harm, as well as the views of their relatives. Our findings relating to clinical services are comparable to those of the previous review (Taylor et al., 2009) describing negative attitudes towards organisational barriers and clinical management. This suggests little systemic change in clinical service provision for self-harm in the last 16 years. However, our review also included views on non-clinical services, where staff attitudes and therapeutic contact were experienced more positively than in clinical settings.

Patients and relatives reported a lack of individualised and collaborative care within clinical services. This was characterised by superficial and formulaic contact that failed to recognise the complexity of self-harm presentations. These findings may be underpinned by the use of increasingly manualised approach within clinical settings as a means of managing high service demands (Hawton, Lascelles, Pitman, Gilbert, & Silverman, 2022). Clinical staff themselves have previously reported conflicts between meeting professional regulations and providing holistic care (Bhui, 2016).

Our review highlighted that genuine and sensitive therapeutic contact in clinical and non-clinical services was viewed as a positive experience that patients linked to promoting recovery, a finding which comes as no surprise. Previous research has shown how strong therapeutic rapport enables patients to feel valued and acknowledged, leading to increased self-esteem and reduced self-harm ideation (Berg et al., 2020; Elliott, Colangelo, & Gelles, 2005). One reason why efforts to establish strong therapeutic rapport are not apparently occurring as standard is the stigmatising beliefs held by some mental health professionals that were also described in our review. Previous research examining staff attitudes in EDs, inpatient and primary care services have revealed stigmatising beliefs, mistrust in patients, and reduced compassion toward people who self-harm (MacDonald et al., 2020; Rayner, Blackburn, Edward, Stephenson, & Ousey, 2019; Saunders, Hawton, Fortune, & Farrell, 2012; Vistorte et al., 2018). This difference in attitudes between services may be attributed to a lack of mental health training for staff in primary care, EDs, and other clinical services not traditionally developed for frontline mental healthcare (Caulfield, Vatansever, Lambert, & Van Bortel, 2019). Our findings demonstrate the importance of specialised training around self-harm, to instil positive attitudes and compassion in clinical staff (Ferguson et al., 2019).

The review also highlighted practical difficulties across services pertaining to waiting times, access, and understaffed services. As this finding is comparable to the findings of the previous systematic review (Taylor et al., 2009), it suggests that there has been no tangible investment or improvement in ED services over that period. High service demands are another potential explanation for the rushed and superficial care reported. There has been a large increase in self-harm presentations, especially by adolescents, in recent years putting further pressures on services (Gunnell et al., 2020; McManus et al., 2019). Previous research has highlighted how overwhelmed staff lack the time and resources to provide effective care (Baker & Naidu, 2021; Mahony, 2014).

Perspective on our findings was provided by an individual with lived experience of accessing self-harm services, which is provided to complement our discussion (supplementary materials: S2). Their perspective is that developments in service provisions over the past 15 years have led to exclusion of those who self-harm, and there is no (or limited) long-term treatment offered to people who self-harm. Psychosocial assessments are often seen as a ‘tick-box’ exercise and do not lead to a concrete treatment plan. They suggest that people with lived experience of self-harm should co-produce training for mental health professionals that is trauma-informed and reduces stigma, particularly for those with personality disorders.

### Limitations

Our quality assessment highlighted four studies of low to moderate quality (Bantjes et al., 2017; Cross & Clarke, 2022; Cully et al., 2022; Mughal et al., 2021), but we included these with equal weighting to other studies in our synthesis for comprehensiveness. However, we acknowledge that these may potentially introduce bias. We limited our initial search to studies published in English, which may explain why all included studies were published in high-and middle-income countries. Moreover, only three of the included studies provided information on participant ethnicity, having either a majority or only white-Caucasian participants. Research has demonstrated that Black, Asian, and Minority Ethnic (BAME) groups experience poor access and quality of care from services due to poor cultural sensitivity and discrimination (Al-Sharifi, Krynicki, & Upthegrove, 2015; Memon et al., 2016). Important attitudes from BAME groups may not have been captured in this review. Furthermore, we did not present findings by age group. This is important considering that self-harm is most prevalent in young people, with both young people and older adults demonstrating high levels of undisclosed self-harm and reduced help-seeking (Gillies et al., 2018; Memon et al., 2016; Troya et al., 2019). Different services are also available for different age groups (e.g., child and adolescent services or adult services), leading to potentially different attitudes.

Included studies inconsistently reported on patients’ histories of self-harm and clinical management. Therefore, we could not interpret findings in the wider context of patients’ previous experiences of services. Similarly, none of the included studies explicitly examined level of suicidal ideation, and the studies examining both attempted suicide and self-harm presentations did not differentiate findings between the two. While in the UK it is customary not to distinguish between episodes on the basis of intent (Kapur, Cooper, O’Connor, & Hawton, 2013), it is possible that one-off or frequent attendance for recurrent ‘non-suicidal self-harm’ elicits a less intense service response than presentations where suicidal intent is expressed, creating different experiences of care. As included studies did not permit us to examine this, there is a need for further research examining how experiences differ by suicidal intent.

### Implications

Our findings show that attitudes towards clinical services have shown little improvement in the 16 years since the previous review by a UK-based team (Taylor et al., 2009). This suggests that the range of UK-based (Health, 2017; NICE, 2013) and international (World Health, 2014) guidelines and policies designed to support service provision have had limited impact. To drive real progress in service provision it may be useful to review guidelines based on these findings. Furthermore, the problems commonly identified by patients (long waiting times, understaffing and limited access to services) have clear implications for the expansion of services, which should be a priority for governments internationally.

With negative staff interactions having a major impact on patient attitudes, policymakers must consider recommendations previously made regarding effective staff training within clinical services (Taylor et al., 2009). Improving staff attitudes and knowledge has been shown to have a wide-scale impact on service quality (Ferguson et al., 2019). This, in turn, has the potential to improve the therapeutic value of psychosocial assessment and improve outcomes demands (Hawton et al., 2022). It may also reduce costs and pressure on services (N. Kapur et al., 2013). Our review also highlighted problems with staff interactions viewed as too standardised and superficial. This demonstrates the importance of the therapeutic relationship, whereby staff should build strong rapport with patients and relatives, involve them in treatment decisions and encompass sufficient flexibility in treatments to ensure that practice is person-centred.

This review substantiates the need for integrated services to maintain quality of care during therapeutic contact, discharge, and transitions in treatment. This is of particular importance during repeated service redesign, especially throughout periods in which the COVID-19 pandemic has impacted service provision. With transformations in services and diversions away from EDs towards other primary, community-based, and remote treatments, including mental health crisis hubs, better collaboration between services can promote effective care while reducing service pressure.

### Future research

With findings demonstrating little improvement in clinical services in the last 16 years, health service researchers and policymakers should monitor the implementation of service guidelines. Research should also address the large gap in the literature pertaining to the attitudes of under-represented groups including older adults, BAME communities and those from low-and middle-income countries. Such groups can offer vital insights that may have not yet been uncovered to broaden our understanding of the quality-of-service provision. Finally, research should evaluate the impact of training and specific service changes on patients and carers’ perceptions of services.

### Conclusions

The findings of this review provide insights into attitudes of individuals who self-harm and their relatives toward clinical and non-clinical services, which remain largely unchanged since a previous review 16 years ago. Across services, experiences of organisational and clinical management were largely negative, while staff attitudes and therapeutic contact were more positively experienced in non-clinical services compared to clinical services. Our findings have important implications for staff training and practice and should be used to reform existing healthcare guidelines for acceptable care for patients who self-harm.

## Supporting information

Supplementary material 1_Search terms

Supplementary material 2_Lived experience commentary

## Financial statement

AP and SR are supported by the National Institute for Health Research (NIHR) University College London Hospital (UCLH) Biomedical Research Centre (BRC).

## Ethical approval

All data are in the public domain.

https://www.cambridge.org/core/journals/psychological-medicine/information/author-instructions/preparing-your-materials

## Data Availability

All data are in the public domain

## Acknowledgments

Many thanks to our lived experience advisor for your insightful perspectives, time and drive to improve the care for people who self-harm.

