## Supplementary material 1_Search terms for "Attitudes towards clinical and non-clinical services among individuals who self-harm or attempt suicide: A systematic review"

**Supplementary material 1 (S1)**

Search terms used in databases

exp Self-injurious Behavior/ OR exp Self-destructive Behavior/ OR exp Self-mutilation/ OR OR exp Self-inflicted wounds/ OR exp Automutilation/ OR exp Suicide, Attempted/ OR exp Suicide/ OR exp Drug Overdoses/ OR (self-harm* OR self?harm* OR self-injur* OR self?injur* OR self-inflict* OR self?inflict OR self-mutilat* OR self?mutilat* OR self-destruct* OR self?destruct* OR self-poison* OR self?poison* OR overdos* OR self-immolat* OR self?immolat* OR auto-mutilat* OR auto?mutilat* OR suicid* OR parasuicid*OR nonsuicid* OR non?suicid* OR NSSI).[tw]

AND

exp Ethnology/ OR exp Ethnography/ OR exp Focus Groups/ OR exp Grounded Theory/ OR exp Phenomenology/ OR exp Observation Methods/ OR exp Questionnaires/ OR exp Health Surveys/ OR (ethnon* OR emic OR etic OR ethnograph* OR participant obser* OR focus group* OR phenomolog* OR grounded theory OR narrative analys?s OR thematic analys?s OR discourse analys?s OR content analys?s OR social constructi* OR semi-structured OR group interview* OR health survey* OR health care survey* OR health?care survey* OR questionnaire*).[tw]

AND

exp Client Attitudes/ OR exp Attitudes/ OR (patient* attitude* OR service user attitude* OR client* attitude* OR service user experienc* OR patient* experienc* OR client* experienc* OR service user perspective* OR patient* perspective* OR client* perspective*).[tw.] OR (famil* OR carer* OR parent*).[tw]
