## Supplementary material 2_Lived experience commentary for "Attitudes towards clinical and non-clinical services among individuals who self-harm or attempt suicide: A systematic review"

**Supplementary material 2 (S2)**

**Lived experience commentary**

I agree completely that negative experiences of services can perpetrate a vicious cycle of self-harm. My experience has often been further episodes due to a wish for corrective experiences from healthcare services. The self-harm has then become a compulsion which is received even more critically by those services due to repetition.

It is important to emphasise that negative experiences of service responses are based on reality. Too often if patients, particularly with a ‘personality disorder’ (PD) diagnosis, report a bad experience, it is put down to our perceptions or dysfunction. If we complain at best, we are told: “we are sorry if you felt that….” There is never an admission that we could be telling the truth.

I think the study should make clear the distinction between physical treatment and psychiatric care. My experience is that A&E department staff have been far more compassionate than mental health services. Psychiatric Liaison teams appear to have a function of deterring attendance while offering no follow up. This can feel like a punitive response which then can lead to more self-harm. The response makes you feel as though you are not worth treating. You then feel it is not worth caring for your body or trying to turn things around.

Often there are no services for self-harm at all. Secondary care mental health services have high access thresholds. Recurrent self-harm may not be treated where patients have been discharged by community services years ago. Patients who self-harm or express suicidal ideation often attract a bad reputation which means they become untouchable by health services. If they present for treatment, they hear: “It is you again”. The patient is blamed for being ‘too dependent’, ‘personality disordered’ and not overcoming ‘dysfunctional behaviour.’

While the study refers to treatment adherence, many patients will not be offered any treatment. Patients can be excluded from IAPT (Increasing Access to Psychological Therapy) services which do not deal with patients at risk of harming themselves. Since patients require psychological interventions there may not be long-term psychological services available in their area and there would be waiting lists.

Many patients who self-harm are given a diagnosis of Borderline Personality Disorder (BPD) with no exploration of their personal circumstances or intervention apart from referral back to the GP (general practitioner). There are PD service pathways, but these may have fixed-term group therapies which patients may not wish to engage with due to personal preference, work commitments or stigma.

Psychiatric Liaison services do not offer treatment, only a one-off chat and signposting to other services. I would argue that these should not be classed as ‘self-harm services’ since they cover all mental health presentations to A&E.

In my own experience, developments in service provision in the last 15 years have led to exclusion of those who self-harm recurrently. In my local hospital there used to be a DSH (deliberate self-harm) team with a social worker and CPN (community psychiatric nurse). This was replaced by a Psychiatric Liaison team which only assesses self-harm as a one off, writes an action plan and signposts. There is no offered treatment or intervention only a diagnosis of BPD.

The study refers to international perspectives. It should be understood that these may not reflect the state of services in the UK. Different countries will vary in their healthcare provision, who is eligible, how it is funded and government regime. It is difficult to bring these countries together in one paper without fully acknowledging their diversity. I cannot imagine self-harm in China which is a tightly controlled state. Suicide in some countries may meet with social rejection. There is such a range in treatment settings, some of which would have been for those who are suicidal rather than self-harming with no intent to end their lives. It is helpful that the studies are summarised to highlight these differences in approach.

There is a difference between self-harm and suicide attempt. A suicide attempt sometimes, though not always, elicits a more concerned response from healthcare services. By contrast self-harm is seen as repetitive, willful, impulsive, and problematic to services.

The study unintentionally highlights the lack of research into self-harm without suicidal involvement. There are useful findings applicable to services and self-harm here, the Australian example about distress in the ED (Emergency Department) is familiar. Several of the UK research studies are very recent and important to the debate.

I thought the findings section was where most insight was shared. There was thought given to what could be learned from research rather than a dry reporting of key messages. The researcher made deep reflections.

The findings about staff attitudes are very strong. Personally, I would use the word ‘disdain’ rather than stigma. Patients who self-harm are seen as time wasting and dysfunctional as though they do not have an actual mental illness. There is a hierarchy of diagnoses in psychiatry and PD is at the bottom. Psychiatrists would avoid PD if they could and often do. I agree entirely that risk assessment does not apply to PD in many cases, since patients are seen to have capacity to make ‘poor decisions’. I also believe that stigma is not a correct word where active prejudice may occur at times.

Belittling attitudes are familiar. I compare it to anorexia if a patient was told she was not thin enough. Patients are often told their injuries are only scratches or superficial. There is no regard or interest to the emotional turmoil which led to that harm. The minimisation then leads to escalation. I do not believe that services do respond to more serious self-harm either since this can still be put down to PD and ‘impulsive’ behaviour. I do not agree that lack of knowledge is the cause given Liaison psychiatrists can minimise self-harm despite having trained for years before choosing to specialise.

I agree that psychosocial assessments are no more than chats with tick boxes which lead nowhere. There is no treatment planning when you are just sent home with nothing. These encounters can become counterproductive and lead to further self-harm. There is a need to review current formats for these assessments so that they may be made more meaningful based on risk assessment, care planning and patient concerns.

Many people with a PD diagnosis are refused medication and told there is nothing wrong with them. This can feel like another form of invalidation and clinical insult. Self-harm could then be a response to that further minimisation of distress.

The main findings of the study put the patient back at the heart of research and improvement in clinical practice. I agree entirely with person-centred care. There is more work to be done though in challenging prejudice, so that it is not acceptable to belittle distress serious enough to result in self-injury. Reflections on the Covid pandemic are important too.

While therapeutic relationships are central to recovery, they are rare in a system that discourages ‘dependency’. Short-term interventions against the recovery model are the norm. Discharge is rapid from secondary care services. GPs manage patients in life threatening situations despite being time pushed themselves.

Service users and survivors with experience of self-harm should coproduce training which should be outside a framework of PD. Training should though be trauma informed to ensure further re-traumatisation is avoided. Training for ED staff is currently led by Psychiatric Liaison teams which may reinforce stigma around patients with PD labels who ‘should be kept out of hospital.’ There is so much blame and shame attached to self-harm and PD that it often feels like a hopeless area to reform attitudes. To start with we should improve the language used, avoiding terms such as maladaptive, dysfunctional and PD. There should be respect towards a patient who self-harms or attempts suicide, since they are a patient and in desperate need.

It can be more difficult to access self-harm care in this age of integration. 111 First means you need to call 111 before going to A&E. You have to wait an hour for a call back from a validation clinician by which time you may have become too afraid to seek treatment. Primary care does not stitch wounds or treat overdoses.

I do respectfully disagree that there are expanding options for the management of self-harm. Statutory services give out the Samaritans number despite this not being dedicated to self-harm. The Samaritans or Mind cannot treat a wound or overdose. People with serious distress need healthcare services. Management of self-harm needs healthcare professionals and not offloading to voluntary sector services without safe systems. I would recommend Self-Injury Support as an excellent charity service. But like many charities, it will struggle with funding and operate a limited service compared with the overwhelming need.
